# Efficacy, safety, and lot to lot immunogenicity of an inactivated SARS-CoV-2 vaccine (BBV152): a, double-blind, randomised, controlled phase 3 trial

**DOI:** 10.1101/2021.06.30.21259439

**Authors:** Raches Ella, Siddarth Reddy, William Blackwelder, Varsha Potdar, Pragya Yadav, Vamshi Sarangi, Vinay Kumar Aileni, Suman Kanungo, Sanjay Rai, Prabhakar Reddy, Savitha Verma, Chandramani Singh, Sagar Redkar, Satyajit Mohapatra, Anil Pandey, Pajanivel Ranganadin, Raghavendra Gumashta, Manish Multani, Shameem Mohammad, Parul Bhatt, Laxmi Kumari, Gajanan Sapkal, Nivedita Gupta, Priya Abraham, Samiran Panda, Sai Prasad, Balram Bhargava, Krishna Ella, Krishna Mohan Vadrevu, the COVAXIN Study Group

## Abstract

**Background:** We report the clinical efficacy against COVID-19 infection of BBV152, a whole-virion inactivated SARS-CoV-2 vaccine formulated with a Toll-like receptor 7/8 agonist molecule adsorbed to alum (Algel-IMDG).

**Methods:** We did a double-blind, randomised, multicentre, phase 3 clinical trial in 25 Indian hospitals to evaluate the efficacy, safety, and immunological lot consistency of BBV152. Healthy adults (age 18–98 years) randomised 1:1 using a computer-generated randomisation scheme received two intramuscular doses of vaccine or placebo administered four weeks apart. The primary outcome was laboratory-confirmed symptomatic COVID-19, occurring at least 14 days after the second dose. Secondary outcomes were efficacy in sub-groups for age (18–< 60 years and ≥ 60 years) and in participants with pre-existing stable medical conditions. We also evaluated safety, reactogenicity, and consistency of immune responses for three consecutive manufacturing lots.

**Findings:** Between November 16, 2020 and January 7, 2021 we recruited 25,798 participants who were randomised to BBV152 or placebo groups; 24,419 received two doses of BBV152 (n = 12,221) or placebo (n = 12,198). In a case-driven analysis, 130 cases of symptomatic COVID-19 were reported in 16,973 (0·77%) participants with follow-up at least two weeks after the second vaccination; 24 occurred in the vaccine group and 106 in placebo recipients giving an overall vaccine efficacy of 77·8% (95% CI: 65·2–86·4). Sixteen cases, one vaccinee and 15 placebo recipients, met the severe symptomatic COVID-19 case definition giving a vaccine efficacy of 93·4% (57·1–99·8). Efficacy against asymptomatic COVID-19 was 63·6% (29·0–82·4). BBV152 conferred 65·2% (95% CI: 33·1–83·0) protection against the SARS-CoV-2 Variant of Concern, B.1.617.2 (Delta). BBV152 was well tolerated with no clinically or statistically significant differences in the distributions of solicited, unsolicited, or serious adverse events between vaccine and placebo groups. No cases of anaphylaxis or vaccine-related deaths were reported.

**Interpretation:** BBV152 was immunogenic and highly efficacious against symptomatic and asymptomatic COVID-19 variant associated disease, particularly against severe disease in adults. Vaccination was well tolerated with an overall incidence of adverse events observed over a median of 146 days that was lower than that observed with other COVID-19 vaccines.

**Funding:** This work was supported and funded by Bharat Biotech International Limited and partly co-funded by the Indian Council of Medical Research.

Clinicaltrials.gov: NCT04641481

## INTRODUCTION

Severe acute respiratory syndrome coronavirus 2 (SARS-CoV-2), a novel human coronavirus, has spread globally causing the COVID-19 pandemic [1]. Vaccines from multiple manufacturers are needed to address the global demand for SARS-CoV-2 vaccines as there is currently insufficient supply. Furthermore, the widely publicised mRNA-based and viral vector vaccines that have been shown to be effective themselves introduce cold chain hurdles and vaccine wastage making them difficult to adopt for many countries.

Bharat Biotech has developed BBV152, a COVID-19 vaccine based on the whole-virion SARS-CoV-2 vaccine strain NIV-2020-770 inactivated with ß-propiolactone. Preclinical studies in rodents and nonhuman primates (NHP) have demonstrated appropriate tolerability, immune responses and protective efficacy [2–4]. We previously reported interim findings from phase 1 and 2 controlled, randomised, double-blind trials on the safety, reactogenicity and immunogenicity of different formulations, which resulted in the selection of a formulation containing a 6 µg dose formulated with a Toll-like receptor 7/8 agonist molecule adsorbed to alum (Algel-IMDG) for further clinical development [5,6]. In use, BBV152 is stored between 2°C and 8°C, which will ease immunisation cold chain requirements. Here, we report findings from a phase 3 case-driven efficacy study including a sub-set analysis of efficacy against newly identified variants of SARS-CoV-2. We also present a nested controlled, randomised, double-blind trial on the safety and immunogenicity of the selected BBV152 formulation, including comparisons of immune responses to three consecutive manufacturing lots measured at day 56, one month after the second dose.

## METHODS

### Study Design and Participants

We assessed the efficacy, safety and immunogenicity of two intramuscular 6 µg Algel-IMDG doses of BBV152 in a randomised, blinded, placebo-controlled, multi-centre study done in 25 centres in India. The trial was approved by the National Regulatory Authority (India) and the respective Ethics Committees of each study centre and was conducted in compliance with all International Conference for Harmonization (ICH) Good Clinical Practice guidelines. The trial was registered on clinicaltrials.gov: NCT04641481.

Participants were adult volunteers 18 years of age or older who were healthy or had stable chronic medical conditions. Volunteers were screened for eligibility based on their health status, including their medical history, vital signs, and physical examination results. Eligible participants provided signed and dated informed consent forms at enrolment. Key exclusion criteria included any diagnosis with an immunocompromising condition, or treatment with immunosuppressive therapy. Detailed inclusion and exclusion criteria can be found in the Protocol (*Supplementary appendix 2)*. A minimum of 20% of the entire sample size was to be comprised of “at-risk participants” defined as being either over 60 years of age, having a coexisting comorbidity (cardio-vascular, diabetes, or any other chronic stable condition), or having a BMI ≥ 35 kg/m^2^. A maximum of 5% of the total enrolled participants were selected from members of the healthcare community.

The primary study objective was to assess the efficacy of the study vaccine in preventing PCR-confirmed symptomatic COVID-19 in a case-driven manner, together with sub-group analyses of asymptomatic efficacy and symptomatic efficacy according to age (18–59 and ≥ 60 years of age), and any chronic stable, medical condition. Major secondary objectives were assessments of the safety and immunogenicity of BBV152 in sub-groups of participants.

### Randomisation and masking

Unblinded statisticians (Cytespace Research and Octalsoft) were involved in designing the randomisation plan and the interactive web response system (IWRS) system for the study. The randomisation plan, stratified for the presence or absence of chronic conditions, was used to generate treatment allocation. The master randomisation list, containing the randomisation number and intended treatment allocation, as well as the kit code, was sent to the IWRS and kits were despatched to the sites according to the IWRS by an unblinded statistician from the CRO tasked with labelling of vaccine vials and the generation of the master randomisation code. Participants were assigned a computer-generated randomisation code and each vial was labelled with a unique code that ensured appropriate masking. The IWRS system assigned the same treatment group for the second visit. Participants, investigators, study coordinators, study-related personnel, and the sponsor were masked to the treatment group allocation, and masked study nurses at each site were responsible for vaccine preparation and administration.

### Procedures

BBV152 (Bharat Biotech, Hyderabad, India) is a whole-virion ß-propiolactone-inactivated SARS-CoV-2 vaccine. The vaccine strain NIV-2020-770 contains the D614G mutation, which is characterised by an aspartic acid to glycine shift at amino acid position 614 of the spike protein [7]. Each 0.5 mL dose contains 6 µg of virus antigen formulated with Algel-IMDG, an imidazoquinoline class molecule that is a Toll-like receptor (TLR) 7/8 agonist (IMDG) adsorbed to Algel. Placebo vials contained the Algel formulation alone without IMDG or inactivated virus antigen. Vaccine and placebo were supplied and stored in a single-use glass vials at 2°C to 8°C, with no on-site dose preparation necessary. The appearance, colour, and viscosity were identical for vaccine and placebo.

At the screening/vaccination visit (visit 1), participants were evaluated with both SARS-CoV-2 reverse-transcriptase–polymerase-chain-reaction (PCR) (ICMR-NIV 2019 nCOV Assay Kit V 3.1) and serology tests (Merilisa, ICMR-NIV Anti-SARS CoV-2 Human IgG ELISA COVID KAVACH), before each injection (*Supplementary materials, pages 5–6)*. Regardless of the outcome of these tests, participants were randomly allocated using the IWRS in a 1:1 ratio to receive two doses of vaccine or placebo on days 0 and 28. Participants who were subsequently found to have a positive PCR test were excluded from receiving the second dose. All females had a urine pregnancy test.

Participants were monitored for 2 hours after vaccination for any acute reactions. No prophylactic medication (ibuprofen/acetaminophen) was prescribed either before or after vaccination. Participants were instructed to record local and systemic reactions daily for seven days after each vaccination (days 0 to 7 and days 28 to 35) using a paper-based memory aid which solicited local and systemic adverse events. Solicited local adverse events included pain at the injection site and swelling, and systemic adverse events included fever, fatigue/malaise, myalgia, body aches, headache, nausea/vomiting, anorexia, chills, generalised rash, and diarrhoea. The memory aid contained fields for symptom onset, severity, time to resolution, and concomitant medications and was collected during the subsequent visit to the site. Routine telephone calls were scheduled following the first seven days after each vaccination. Participants reported all unsolicited adverse events and serious adverse events throughout the study. Adverse events were graded according to severity (mild, moderate, or severe) and by relationship (related or unrelated) to the investigational vaccine, as detailed in the protocol.

Study sites were classified into three categories: Category 1: in addition to administering the vaccine or placebo, a series of post-dose follow-up telephone calls (every two weeks) were scheduled to detect suspected symptomatic COVID-19 (n = 16,477) and those who met symptomatic criteria had a clinical assessment (*Protocol, Supplementary appendix 2*), and a nasopharyngeal swab (NP) was taken for PCR confirmation. Category 2: in addition to symptomatic follow-up, a series of post-dose 2 NP swabs were collected on-site for detection of asymptomatic COVID-19 infection at monthly intervals (n = 8,721); Category 3: in addition to follow-up for symptomatic and asymptomatic COVID-19 infection, blood samples were collected for immunological assessments (n = 600). Unscheduled illness visits were encouraged for participants till day 360 (± 14 days). All participants were instructed to contact the team on an as-needed basis.

### Outcomes

The primary outcome was the efficacy of the BBV152 vaccine in preventing a first occurrence of symptomatic COVID-19 (any severity) with onset at least 14 days after the second dose in the per-protocol population composed of participants who were SARS-CoV-2 negative by PCR and serology at baseline, had no major protocol deviations, and followed-up for at least two weeks after the second dose. End points were judged by an independent adjudication committee masked to treatment allocation. COVID-19 cases were defined as participants with at least two of the following symptoms: fever (temperature ≥ 38°C), chills, myalgia, headache, sore throat, or new olfactory or taste disorder, or had at least one respiratory sign or symptom (including cough, shortness of breath, or clinical or radiographic evidence of pneumonia) and at least one SARS-CoV-2 PCR-positive nasopharyngeal swab. COVID-19 cases were followed daily to assess symptom severity until symptoms resolved. In PCR-positive participants who consented, an additional NP swab for genotyping and a blood sample for evaluating correlates of protection were collected. Secondary efficacy outcomes included efficacy in subgroups defined by age (18–59 years and ≥ 60 years), gender, and health risk for severe disease (presence or absence of a coexisting chronic medical condition), efficacy against variants of concern, and efficacy against asymptomatic infections occurring after receipt of two doses of vaccine/placebo periodically at a month’s interval in 8,721 participants, among whom 6,289 participants were SARS-CoV-2 negative at baseline and included in the per protocol analysis.

The immunological secondary outcome was evaluation of consistency of immune responses from three consecutive manufacturing lots. This was based on geometric titres (GMTs) evaluated using a wild-type virus microneutralisation assay (MNT_50_) (*Supplementary materials, page 8*). Immune responses against three SARS-CoV-2 epitopes, the S1 protein and the receptor binding domain (RBD) of the spike protein, and the nucleocapsid antigen (N-antigen) were measured as IgG responses by ELISA (*Supplementary materials, page 9*). All sera were analysed in a blinded manner at Bharat Biotech (Hyderabad, India) and submitted to the CRO for data analysis and preparation of the report. Safety secondary outcomes were the proportions of participants with solicited local and systemic reactogenicity within seven days after vaccination, and with unsolicited adverse events recorded within 28 days after vaccination.

### Statistical Analysis

The study was designed to obtain a two-sided 95% CI for vaccine efficacy with lower bound ≥ 30%. Based on a true efficacy of 60% and power of 85%, the case-driven trial was planned to accrue 130 cases. Assuming 1% incidence of PCR-confirmed symptomatic COVID-19 disease among placebo recipients during follow-up beginning 14 days after the second dose, the number of participants required to accrue 130 cases was approximately 18,572. To allow for a 20% baseline seropositivity rate or PCR-confirmed COVID-19 and 10% loss to follow-up, we planned to enrol 25,800 participants. Sample size estimation was performed using PASS 13 software (NCSS, Kaysville, Utah, USA).

Estimation of vaccine efficacy was based on person-time incidence rates: VE = 1 – (nv/Fv) / (np/Fp) = 1 – R, where R = (nv/Fv) / (np/Fp); nv and np are the numbers of participants who develop PCR-confirmed symptomatic COVID-19 among BBV152 vaccine and placebo recipients, respectively, and Fv and Fp are the corresponding total lengths of follow-up in years in the two groups, with follow-up in years defined as follow-up in days divided by 365.25. We also define the parameter P, the proportion of participants with COVID-19 who were in the vaccine group. Then a two-sided confidence interval (CI) around the estimated VE is obtained by converting an exact CI for the probability parameter P, using the observed Fp/Fv, to a CI for VE. Interim analyses were planned at 43 and 87 primary endpoint cases, using an O’Brien-like Lan-DeMets alpha spending function [8].

Safety endpoints are reported as number and % of participants. Immunological endpoints are expressed as GMTs with 95% confidence intervals (CIs) calculated from 95% CIs for means of log_10_ (titre), which used t-distributions. The criterion for consistency (equivalence) (equivalence) of the immune response to BBV152 across three consecutive manufacturing batches was that two-sided 95% CIs for the ratio of GMTs for all pairs of lots be entirely contained within the interval [0.5, 2.0], limits which have frequently been used for the related concept of non-inferiority in vaccine trials [9].

For continuous variables (less than 20 observations), medians and IQRs are reported. Exact binomial calculations were used for the CI estimation of proportions. Wilson’s score test was used to test differences in proportions. A result with two-sided P ≤ 0.05 or one-sided P ≤ 0.025, as appropriate, was considered statistically significant. This report contains results regarding immunogenicity and safety outcomes (captured on days 0 to 56) and efficacy results with a median of 99 days (two weeks after a second dose). Certain prespecified subgroup analyses are not included in this report but will be presented in future analyses when a larger dataset is available. Descriptive and inferential statistics were performed using SAS 9·4.

## RESULTS

Between November 16, 2020 and Jan 7, 2021, we screened 26,028 volunteers and recruited and vaccinated 25,798 participants across 25 sites (**Figure 1**). At the data cut-off date of May 17, 2021, a total of 23,803 (92·3%) participants had a median of 146 days of safety data available after the first dose. Among these participants, 7058 (27·5%) had at least one coexisting condition. The mean age was 40·1 years, and 10·7% of participants were older than 60 years of age. A large proportion of participants were seropositive at baseline (30%) and were thus excluded from the per-protocol analysis but contributed to the safety dataset. All baseline characteristics were similar between vaccine and placebo groups (**Table 1**).

**Table 1:** Demographic of participants in the safety population

| Parameter | BBV152<br>n (%) | Placebo<br>n (%) |
| --- | --- | --- |
| **N =** | 12,879 | 12,874 |
| **Age, years** |  |  |
| (Mean ± SD) | 40·1 ± 13·8 | 40·1 ± 14·1 |
| Range | 18, 92 | 19, 97 |
| **Sex, n (%)** |  |  |
| Female | 4214 (32·7) | 4254 (33·0) |
| Male | 8665 (67·3) | 8620 (66·9) |
| **Body Mass Index (BMI), kg/m<sup>2</sup>** |  |  |
|  | 24·3 ± 4·4 | 24·3 ± 4·3 |
| **Pre-existing medical conditions, n (%)** |  |  |
| Stable cardiovascular disease | 557 (4·3) | 523 (4·1) |
| Stable respiratory disease | 126 (1·0) | 170 (1·3) |
| Controlled diabetes | 706 (5·5) | 735 (5·7) |
| Stable liver disease | 25 (0·2) | 28 (0·2) |
| Severe obesity (BMI > 35) | 56 (0·4) | 94 (0·7) |
| Other stable co-morbidities | 839 (6·5) | 910 (7·1) |
| Multiple risk categories | 458 (3·6) | 497 (3·9) |
| **Baseline assessments for SARS-CoV-2 positivity\*** |  |  |
| Positive for anti-SARS-CoV-2 IgG | 3932 (30·5) | 3886 (30·2) |
| Positive for SARS-CoV-2 by PCR | 108 (0·8) | 105 (0·8) |

\* At the screening or initial vaccination visit (visit 1) participants were evaluated for exposure to SARS-CoV-2 with both anti-SARS-CoV-2 IgG by ELISA and reverse-transcriptase polymerase chain reaction (PCR). Regardless of the outcome of these tests, participants were randomised and allocated to a group.

**Figure 1:**
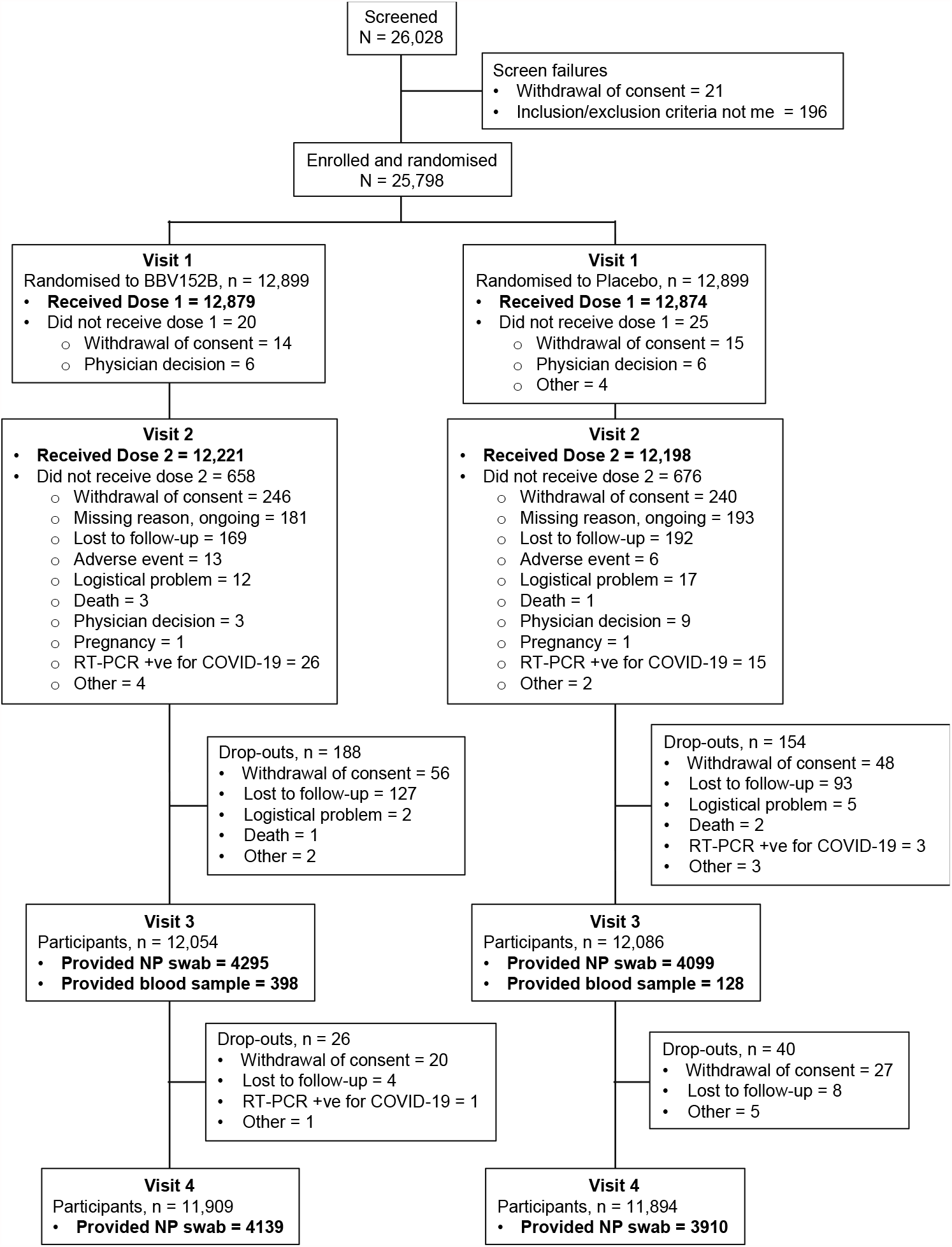
CONSORT Flow Diagram

### Efficacy

Among the 16,973 participants in the per protocol analysis population (*Supplementary table 2, page 10*), the planned efficacy analysis occurred after the accrual of 130 symptomatic COVID-19 cases which started to present soon after the beginning of the observation period (**Figure 2**). There were 24 (0·28%) cases among 8471 participants in the vaccine arm and 106 (1·25%) cases among 8502 participants in the placebo group, resulting in estimated vaccine efficacy of 77·8% (95% CI: 65·2–86·4). There were sixteen cases who met the severe symptomatic COVID-19 cases definition, all but one of whom were in the placebo group, resulting in a vaccine efficacy of 93·4% (95% CI: 57·1–99·8). Efficacy against asymptomatic COVID-19 infections was 63·6% (29·0–82·4). In the 1858 elderly participants in the analysis, the split of cases between vaccine and placebo groups was 5 (0·56%) of 893 participants and 16 (1·66%) of 965, respectively, giving an efficacy of 67·8% (8·0–90·0). Efficacy in the 15,115 participants who were younger than 60 years was 79·4% (66·0–88·2) (**Table 2**).

**Table 2:**
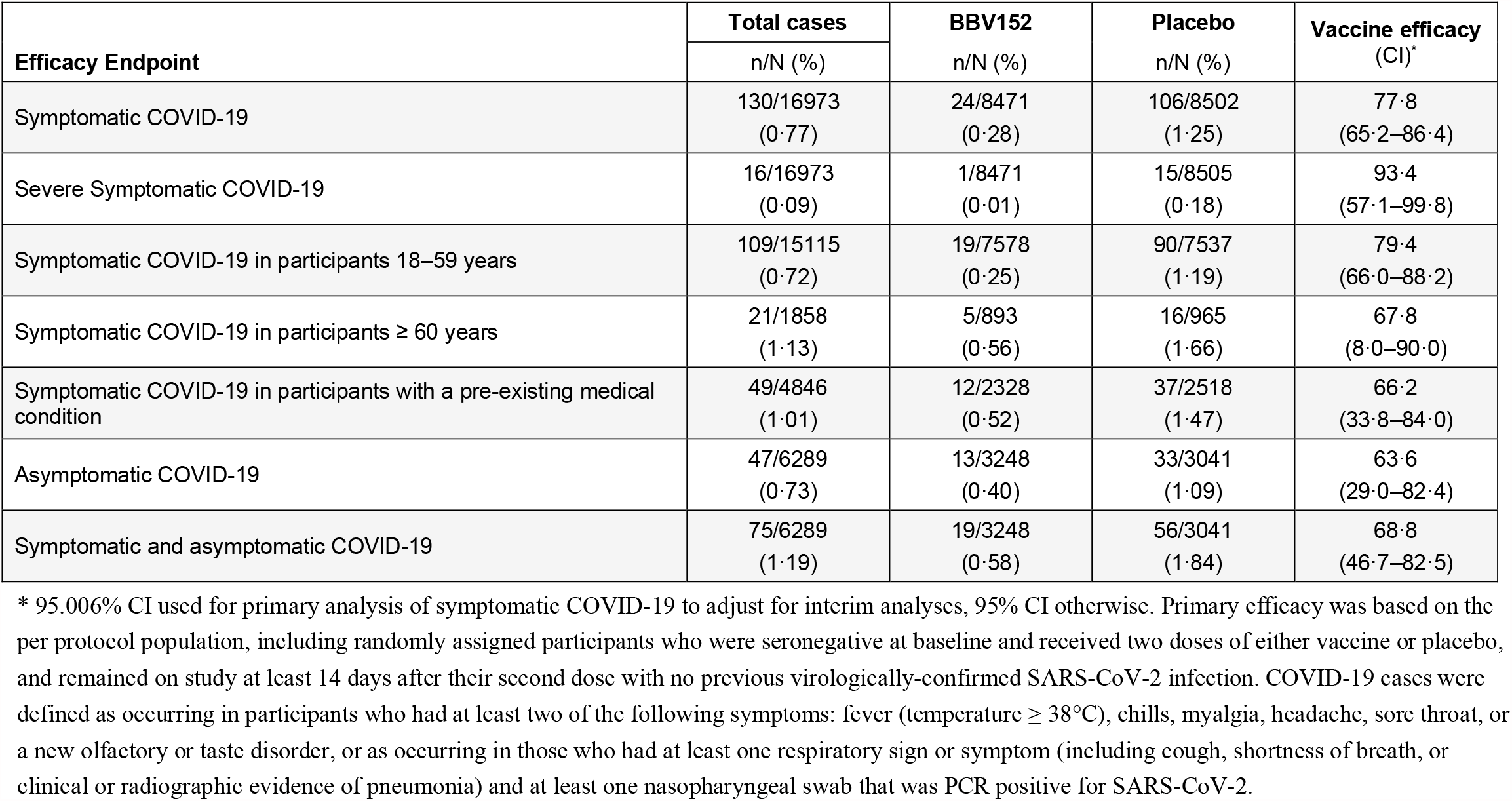
Efficacy against SARS-CoV-2 after at least 14 days following a second dose of BBV152 vaccine in the per protocol population.

**Figure 2:**
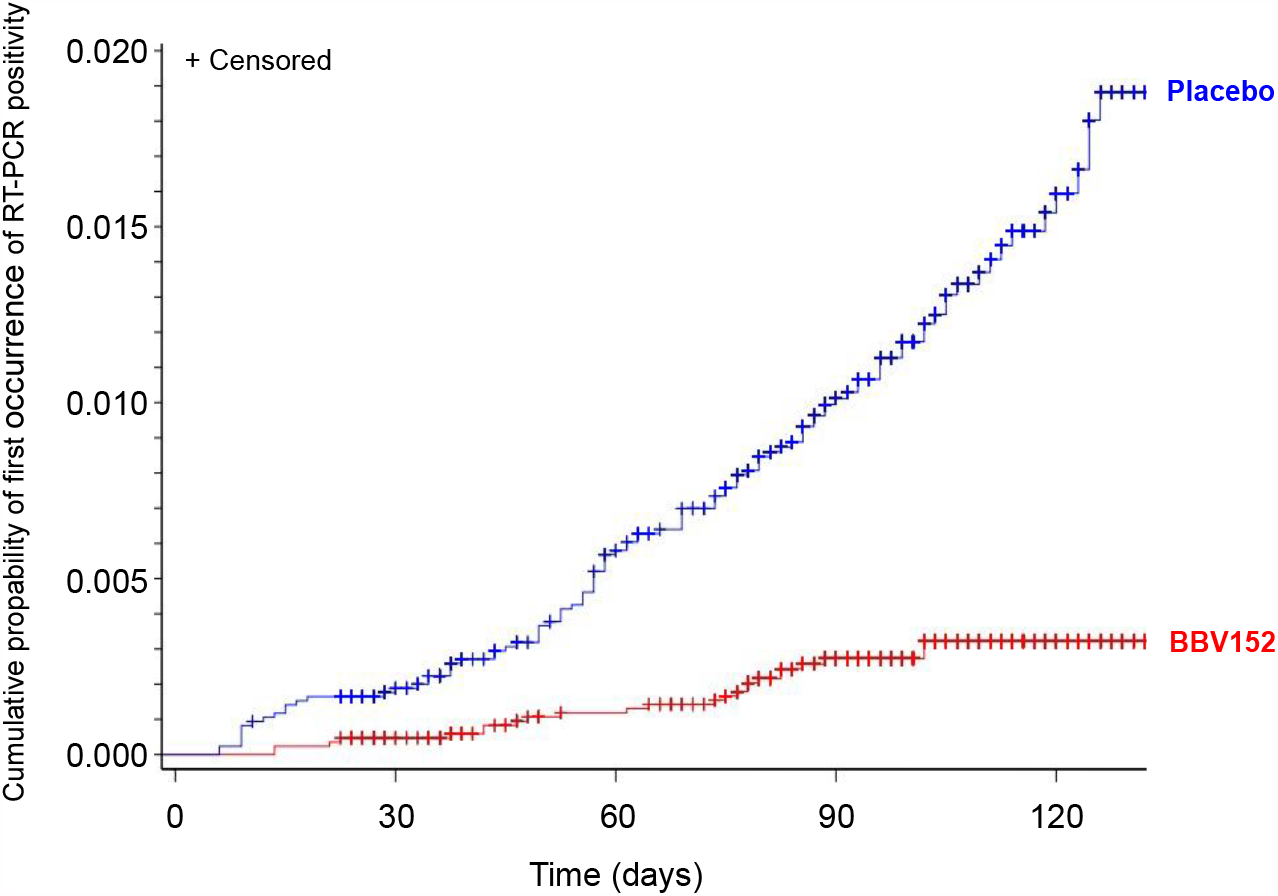
Kaplan Meier plot of first occurrence of virologically confirmed (RT-PCR positive) symptomatic cases of COVID-19 (per-protocol set)

### Immune Responses

At day 56 in the groups who received lots 1, 2, 3 or placebo GMTs (MNT_50_) of SARS-CoV-2 neutralising antibodies were 130·3 (95% CI: 105·8–160·4), 121·2 (97·6–150·5), 125·4 (101·3–155·1), and 13·7 (10·7–17·4), respectively (**Table 4**). GMT ratios between all three pairs of lots were consistently similar: lots 1:2 GMT ratio 1·08 (95% CI: 0·80–1·45), lots 1:3 GMT ratio 1·04 (0·77–1·40), and lots 2:3 GMT ratio 0·97 (0·71–1·31). All the 95% CIs for the GMT ratios were contained within the interval [0·50, 2·0] (*Supplementary figure 1, page 11*), meeting the predefined criterion for a consistent immune response across lots.

There were no marked differences in GMTs for neutralizing antibodies at Day 56 when assessed based on age or gender (*Supplementary table 3, page 12*). The GMT was higher (194.3 [95% CI: 134.4–280.9, n = 48] in vaccinees who were seropositive for SARS-CoV-2 IgG at baseline than in those who were seronegative (118.0 [104.0–134.0]).

**Table 3:**
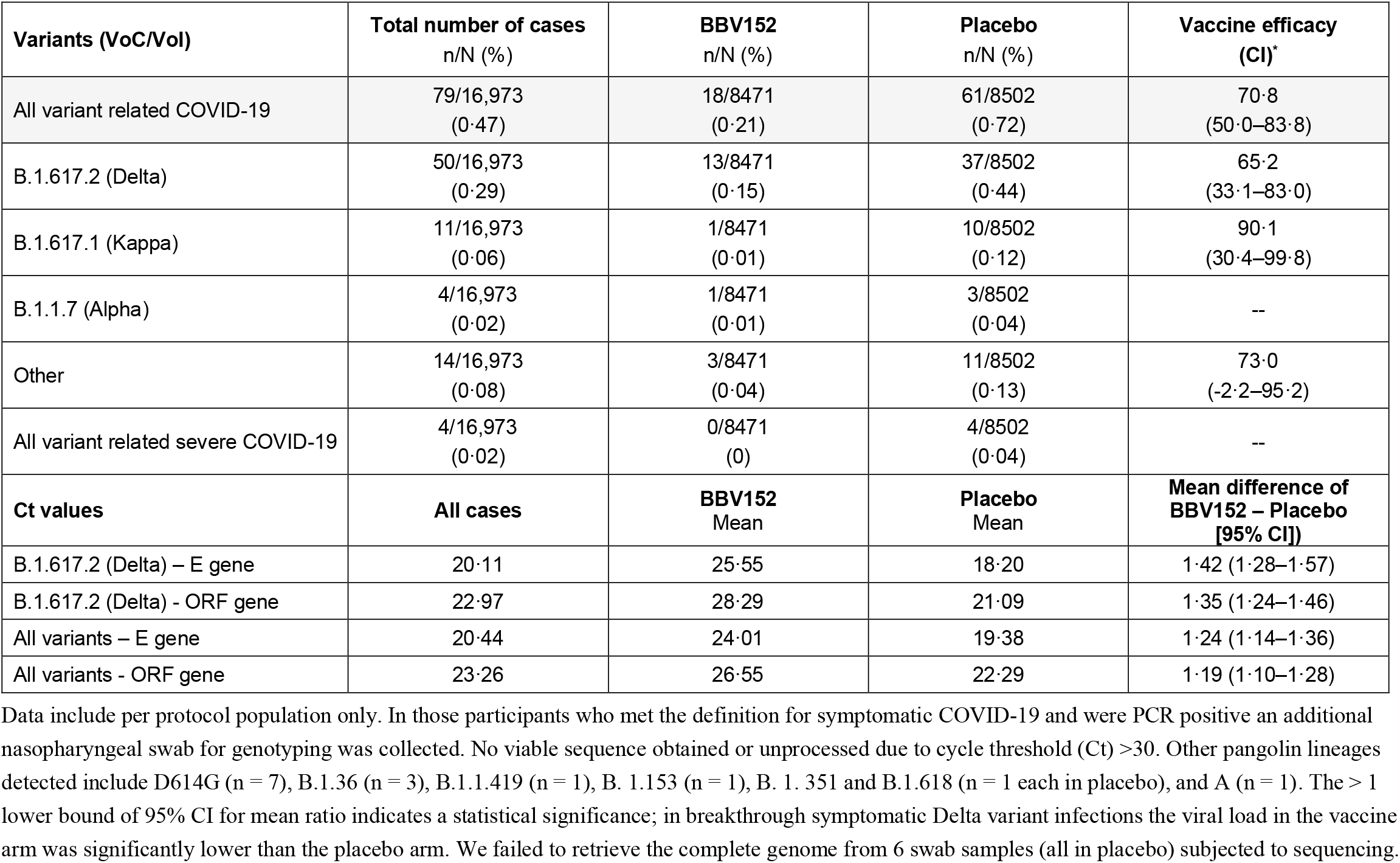
Efficacy against variants of interest (VoI) and concern (VoC).

| **Variants (VoC/VoI)** | **Total number of cases**<br>**n/N (%)** | **BBV152**<br>**n/N (%)** | **Placebo**<br>**n/N (%)** | **Vaccine efficacy**<br>**(CI)\*** |
| --- | --- | --- | --- | --- |
| All variant related COVID-19 | 79/16,973<br>(0.47) | 18/8471<br>(0.21) | 61/8502<br>(0.72) | 70.8<br>(50.0–83.8) |
| B.1.617.2 (Delta) | 50/16,973<br>(0.29) | 13/8471<br>(0.15) | 37/8502<br>(0.44) | 65.2<br>(33.1–83.0) |
| B.1.617.1 (Kappa) | 11/16,973<br>(0.06) | 1/8471<br>(0.01) | 10/8502<br>(0.12) | 90.1<br>(30.4–99.8) |
| B.1.1.7 (Alpha) | 4/16,973<br>(0.02) | 1/8471<br>(0.01) | 3/8502<br>(0.04) | -- |
| Other | 14/16,973<br>(0.08) | 3/8471<br>(0.04) | 11/8502<br>(0.13) | 73.0<br>(-2.2–95.2) |
| All variant related severe COVID-19 | 4/16,973<br>(0.02) | 0/8471<br>(0) | 4/8502<br>(0.04) | -- |
| **Ct values** | **All cases** | **BBV152**<br>**Mean** | **Placebo**<br>**Mean** | **Mean difference of**<br>**BBV152 – Placebo**<br>**[95% CI]** |
| B.1.617.2 (Delta) – E gene | 20.11 | 25.55 | 18.20 | 1.42 (1.28–1.57) |
| B.1.617.2 (Delta) - ORF gene | 22.97 | 28.29 | 21.09 | 1.35 (1.24–1.46) |
| All variants – E gene | 20.44 | 24.01 | 19.38 | 1.24 (1.14–1.36) |
| All variants - ORF gene | 23.26 | 26.55 | 22.29 | 1.19 (1.10–1.28) |

Data include per protocol population only. In those participants who met the definition for symptomatic COVID-19 and were PCR positive an additional nasopharyngeal swab for genotyping was collected. No viable sequence obtained or unprocessed due to cycle threshold (Ct) >30. Other pangolin lineages detected include D614G (n = 7), B.1.36 (n = 3), B.1.1.419 (n = 1), B. 1.153 (n = 1), B. 1. 351 and B.1.618 (n = 1 each in placebo), and A (n = 1). The > 1 lower bound of 95% CI for mean ratio indicates a statistical significance; in breakthrough symptomatic Delta variant infections the viral load in the vaccine arm was significantly lower than the placebo arm. We failed to retrieve the complete genome from 6 swab samples (all in placebo) subjected to sequencing.

At Day 56 IgG titres to all three epitopes (S1 protein, RBD, and N protein) were detected after two doses. For all three lots combined the GMTs at Day 56 were 9742 EU/mL (95% CI: 8949–10606) for S1 protein, 4124 EU/mL (3731–4557) for RBD-competitive binding, and 4161 EU/mL (3736–4633) for SARS-CoV-2 N protein assays (**Table 4**). The placebo group did not display any meaningful change in titres over the course of the study for any of the immune targets.

**Table 4:** SARS-CoV-2 neutralising (MNT_50_) and binding antibody responses (Anti-S1 protein, RBD-binding, and N protein IgG). Data are shown for neutralising response expressed as MNT_50_ at Days 0 (baseline) and Day 56, four weeks after the second vaccination. Day 56 IgG antibody titres are expressed as arbitrary ELISA units, all baseline titres being at the cut-off for the assay (reciprocal of 1:500 dilution).

| Assay |  | Geometric mean titres (95% CI) at Day 56 in sub-sets of the different study groups |  |  |  |  |
| --- | --- | --- | --- | --- | --- | --- |
|  |  | BBV152 |  |  |  | Placebo |
|  |  | Lot 1 | Lot 2 | Lot 3 | All lots |  |
| SARS-CoV-2 MNT<sub>50</sub> | N = | 132 | 129 | 136 | 397 | 125 |
|  | Day 0 | **9.9**<br>(8.3, 11.9) | **8.6**<br>(7.5, 9.9) | **7.9**<br>(7.0, 8.9) | **8.8**<br>(8.0, 9.6) | **8.9**<br>(7.7, 10.4) |
|  | N = | 128 | 125 | 133 | 386 | 119 |
|  | Day 56 | **130.3**<br>(105.8, 160.4) | **121.2**<br>(97.6, 150.5) | **125.4**<br>(101.3, 155.1) | **125.6**<br>(111.2, 141.8) | **13.7**<br>(10.7, 170.4) |
| S-protein binding IgG | N = | 129 | 124 | 134 | 387 | 121 |
|  | Day 56 | **9760**<br>(8483, 11228) | **10404**<br>(8873, 12198) | **9152**<br>(7912, 10586) | **9742**<br>(8949, 10606) | **1528**<br>(1323, 1765) |
| RBD-binding IgG | N = | 129 | 124 | 134 | 387 | 121 |
|  | Day 56 | **4266**<br>(3584, 5079) | **4423**<br>(3669, 5333) | **3740**<br>(3180, 4399) | **4124**<br>(3731, 4557) | **1443**<br>(1261, 1651) |
| N protein binding IgG | N = | 129 | 124 | 134 | 387 | 121 |
|  | Day 56 | **4551**<br>(3800, 5450) | **4183**<br>(3423, 5111) | **3798**<br>(3165, 4558) | **4161**<br>(3736, 4633) | **1485**<br>(1275, 1730) |

### Safety

There were 15 deaths in the study, none of which were considered by the investigators to be related to the vaccine or placebo; six deaths were reported to be related to COVID-19. In BBV152 recipients there were five deaths all due to causes unrelated to vaccination: cerebellar haemorrhage, haemorrhagic stroke, ovarian cancer with metasases, sudden cardiac death, and COVID-19. Ten placebo recipients died, also from unrelated conditions: alcohol overdose, myocardial infarction, cardiac arrest with underlying hypertension, five from COVID 19 and two which remain to be determined. No anaphylactic events were reported.

The vaccine had a good reactogenicity profile with similar rates of solicited, unsolicited, and serious adverse events and adverse events of special interest in vaccine and placebo groups. Serious adverse events occurred in 99 participants; 39 (0·30%) received BBV152 and 60 (0·47%) received placebo (*Supplementary Table 4, page 13*). Two related serious adverse events were reported among BBV152 recipients. Long-term safety monitoring will continue for 1 year after administration of the first dose of BBV152.

Solicited adverse events analyses are provided for all enrolled 25,798 participants (*Supplementary table 5, page 14*). Overall, incidence rates were lower after the second dose than the first, and tended to be slightly higher in the BBV152 group than the placebo group. However, all incidence rates were low, with only 12·4% reporting any solicited AE after vaccine or placebo. Among the local or systemic solicited AEs, only local injection pain was reported with an incidence greater than 1% (*Supplementary table 4, page 13*). Similar proportions of vaccine (3·04%) and placebo (2·78%) groups reported local pain after the first dose, falling to 1·81% and 1·62% after the second dose, respectively. Other local AEs were reported by less than 0.3% of participants in any group after either dose. Solicited systemic AE were reported less frequently, after 2·57% and 1·92% of first doses of vaccine or placebo, respectively. The most frequent solicited systemic AE overall was headache, followed by pyrexia, fatigue and myalgia but at incidences below 1% in both groups. Rates of local and systemic AEs reported in the BBV152 group as mild (11·2%), moderate (0·8%), or severe (0·3%) were comparable to the placebo group (mild [10·8%], moderate [1·1%], and severe [0·4%]). Unsolicited AEs were reported by 1·8% and 1·7% of vaccinees and placebo recipients, respectively. No significant differences were observed between the vaccine and placebo groups, the P value for all comparisons being > 0·05.

## DISCUSSION

We report findings from the phase 3 efficacy, safety and immunogenicity clinical trial of BBV152, a whole-virion inactivated SARS-CoV-2 vaccine. In the final per-protocol analysis, measured 14 days after the second of two doses of BBV152, there was a vaccine efficacy of 77·8% (95% CI: 65·2–86·4) against symptomatic COVID-19 disease, and perhaps more importantly a higher efficacy against severe COVID-19 of 93·4% (57·1–99·8). Thus, cases of severe disease which require hospitalisation and have threatened to overwhelm healthcare facilities will be markedly decreased in fully vaccinated populations Although the study was not powered to definitively assess efficacy in subgroups with different ages, gender, or the presence of pre-existing comorbid conditions, efficacy rates for symptomatic COVID-19 were all high in these sub-groups (>66%) with the lower limits of the respective 95% CIs being above 30% in all cases except for the > 60 years group.

This phase 3 study confirms our earlier observations on the safety and immunogenicity profiles of BBV152 in phase 1 and 2 trials [5,6]. There were no safety concerns raised, no reports of anaphylactic events after BBV152 administration, and all adverse events (solicited, unsolicited, and serious adverse events) were well balanced between BBV152 and placebo groups. One possibly related serious adverse event in the BBV152 group was a case of immune thrombocytic purpura that occurred 39 days after the second dose in a participant who was SARS-CoV-2 seropositive at baseline, which resolved in four days. After any dose, the combined incidence rate of local and systemic adverse events in this study is noticeably better than the rates for other SARS-CoV-2 vaccine platform candidates [10,11], and comparable to the rates for other inactivated SARS-CoV-2 vaccine candidates [12].

When measured as neutralising antibodies, the three consecutive manufacturing lots of vaccine induced consistent humoral immune responses, and when measured as ELISA IgG responses against three SARS-CoV-2 epitopes (S1 and RBD of the spike protein, and the nucleocapsid antigen) antibody titres were similar across all lots (**Table 4**). Further, BBV152 generated comparable neutralising immune responses in participants < 60 and ≥ 60 years of age (*Supplementary table 3, page 12)*; vaccine efficacy was 79·4 % (95% CI: 66·0–88·2) and 67·8% (8·0–90·0) in the younger and older subgroups, respectively (**Table 2**).

The recent surge in SARS-CoV-2 variant strains has raised concerns regarding the efficacy of vaccines against the new Variants of Concern (VoC). Some COVID-19 vaccines, notably Coranavac and ChAdOx1, have been reported to have diminished efficacy against the Gamma (P1) and Beta (B.1.351) variants first isolated in Brazil and South Africa [11,13]. The ChAdOx1 vaccine is reported to have equivalent efficacy against the Alpha (B.1.1.7) variant, which is widely circulating [14]. Effectiveness after two doses of the mRNA-based vaccine, BNT162b2, decreased from 93·4% (95% CI: 90·4–95·5) against B.1.1.7 to 87·9% (78·2–93·2) against B.1.617.2 [15]. With ChAdOx1 effectiveness after two doses decreased from 66.1% (54.0–75.0) against B.1.1.7 to 59.8% (28.9–77.3) against B.1.617.2 [15]. The emergence of VoC occurred during the conduct of our trial, and we obtained additional consent to collect additional NP swabs from RT-PCR-confirmed symptomatic COVID-19 participants. All sequences were generated by the National Institute of Virology, Pune, India using the quantitative approach [16,17]. Controls were checked to ensure no evidence of amplification in the negative tests and that expected RNA quantification was consistent with cycle threshold (Ct) values provided by the testing laboratories. All samples were processed by laboratory staff masked to vaccine allocation. A total of 79 variants were reported from 16,973 samples, 18 in the vaccine and 61 in the placebo group. Among 50 Delta (B.1.617.2) positive-confirmed cases, 13 and 37 participants were in the vaccine and placebo arms, resulting in vaccine efficacy of 65·2% (95% CI: 33·1–83·0). In breakthrough symptomatic Delta variant infections, based on Ct values, the viral load in the vaccine arm was significantly lower than the placebo arm. Efficacy against the Kappa (B.1.617.1) variant was 90·1% (95% CI: 30·4–99·8). No cases of severe variant-related cases of COVID-19 were reported in the vaccinees but four severe cases were reported in the placebo recipients infected with Alpha, Kappa, Delta, and unclassified variants respectively (Table 3). As previously reported BBV152-induced antibodies show no significant decrease in neutralisation activity against the Alpha (B.1.1.7) variant, but demonstrate marginal reductions in neutralisation activity, by 2-, 2-, 3-, and 2.7-fold, respectively, of the B.1.1.28, B.1.617.1, B.1.351 (Gamma), and B.1.617.2 (Delta) variants [18–21].

No licensed SARS-CoV-2 vaccine has reported efficacy against asymptomatic infection in a randomised controlled trial, based on nucleic acid testing, although the mRNA vaccine, BNT162b2, has been associated with decreased asymptomatic SARS-CoV-2 infections in healthcare workers [22]. Several other vaccine studies employed surrogate markers to assess asymptomatic efficacy by periodically collecting serum from trial participants and assessing for anti-SARS-CoV-2-nucelocapsid binding antibody (N antigen) [10]. In this study, a total of 8,721 participants made monthly clinical visits for routine medical check-ups and collection of NP swabs for PCR confirmation of asymptomatic COVID-19. In the per protocol set, 3,248 and 3,041 participants in BBV152 and placebo groups, respectively, were enrolled and as per the cut-off date, up to two months after the second dose, 14 and 33 positive PCR confirmations have been reported in the vaccine and placebo groups, respectively, an efficacy of 66·6 % (95% CI: 23·7–80·4). A study with the ChAdOx1 vaccine found no efficacy (3·8%) against asymptomatic infections, albeit direct comparisons cannot be made as a surrogate serological marker was used [10]. Our findings corroborate well with preclinical protective efficacy studies in hamsters and NHP, which reported lower and upper airway protection against SARS-CoV-2 infection [3,4].

This study has several limitations. Due to the low number of cases reported between doses 1 and 2, we cannot calculate vaccine efficacy after a single dose. This report contains a median safety follow-up of 146 days for all participants, so long-term safety follow-up of BBV152 is required and is currently underway. The data presented on efficacy against variants other than Delta must be considered preliminary as the numbers reported are small. Additional efforts to assess the clinical efficacy of BBV152 against VoC are being planned. The potential establishment of a correlate of protection is not feasible at the time of this report. Finally, this study population lacked ethnic and racial diversity, underscoring the importance of evaluating the efficacy of BBV152 in other populations.

Although the study was designed to vaccinate and follow participants for one year after the second dose, given the nature of the pandemic in India and the emergency use authorization for BBV152, after meeting the pre-defined efficacy success criteria, the DSMB and sponsor decided to unblind those placebo participants who were eligible to receive an approved COVID-19 vaccine. Unblinding in such cohorts was planned only after the accrual of the protocol pre-specified 130 cases, in a phased manner: health care professionals, individuals ≥45 years, followed by those <45 years. Our sample estimations accounted for 20% seropositivity. As we observed baseline seropositivity rates of 30% and due to the unblinding of the health care professionals and elderly individuals (who are eligible for COVID-19 vaccination), the protocol was amended to expand the sample size to 30,800, with 5,000 additional participants now being enrolled in Brazil. This will ensure the study evaluates the efficacy of BBV152 against VoC and provides an opportunity to accrue additional severe COVID-19 cases as well as more racial diversity. This manuscript contains data from the Indian cohort only.

However, this study does have several strengths. The study enrolled participants with ages ranging from 18 to 98 years and found no major differences in immune responses across the broad age groups of under- and over-60 year-olds. Participants considered to be at-risk of acquiring COVID-19 were prioritised, so a total of 2,750 participants were above 60 years of age and 7,065 reported at least one pre-existing medical condition across ages. To ensure generalisability, this study was conducted with participants from diverse geographic locations, enrolling 25,798 participants across 25 hospitals. This is the first trial to report preliminary promising findings on the efficacy against asymptomatic infections and clinical lot-to-lot immunological comparability.

The most common solicited adverse event was pain at the injection site, followed by headache, fatigue, and fever. No severe or life-threatening (Grade 4 and 5) solicited adverse events were reported. Although the study was not powered to find such differences, no meaningful safety differences were observed between the groups. After any dose, the combined incidence rate of local and systemic adverse events in this study is noticeably better than the rates for other SARS-CoV-2 vaccine platform candidates [23–27] and comparable to the rates for other inactivated SARS-CoV-2 vaccine candidates [28,29].

However, other vaccine studies enrolled different populations and employed varying approaches to measure adverse events.

The positive safety, immunogenicity and efficacy results presented here will support regulatory submissions for emergency use authorisation (EUA) which BBV152 (COVAXIN™) has already received in 23 countries. With the inclusion of Vaccine Vial Monitor 7, storage at 2°C–8°C, and a 28-day open-vial policy (limiting open-vial vaccine wastage by 10–25%), the established efficacy of BBV152 against symptomatic and asymptomatic infection will be critical towards mitigating the COVID-19 pandemic.

## Data Availability

Individual participant (de-identified) data will be made available when the trial is complete upon direct request to the corresponding author with an appropriate research proposal. Once such a proposal is approved data will be shared through a secure online platform.

## ACKNOWLEDGEMENTS

We would like to sincerely thank the volunteers, investigators, study coordinators and healthcare workers involved in this study. We express our gratitude to the teams at IQVIA, Cytespace, and Octalsoft who did the trial. Drs. Shashi Kanth Muni, Ashwini Maratha, Yuvraj Jogdand, Amarnath Sapan Kumar Behera, Jagadish Kumar, Bharagav Reddy, Mr. Sunil Kumar, Ms. Aparna Bathu and Ms. Sandya Rani of Bharat Biotech participated in protocol design and clinical trial monitoring. We thank the members of the DSMB and Adjudication Committee for their continued support and guidance of this ongoing clinical study. This vaccine candidate could not have been developed without the efforts of Bharat Biotech’s Manufacturing, and Quality Control teams. We are grateful to Keith Veitch (keithveitch communications, Amsterdam, The Netherlands) for editorial assistance with the manuscript.

## Author Contributions

All authors met the criteria for authorship set forth by the International Committee for Medical Editors. RE contributed to the manuscript preparation and KMV, SPr, KE, WB, NG, SPa, PA, and BB reviewed the manuscript. RE and KMV and were responsible for overall project coordination. SRe, VS, and VA were led clinical operations and helped immensely with designing the protocol. WB was involved with the study design and statistical analysis plan. VP, PY, and GS led the virological confirmation and genomic sequencing efforts. The contract research organisation (IQVIA) was responsible for analysing the data and generating the report. All principal investigators (SK, SR, PRe, SV, CS, SR, SM, AP, PRa, RG, MM, SM, PB, and LK) were involved in the scientific review of this paper. RE, KMV, PY, GS, VA, and VS had full access and verified the masked data in the study, and can vouch for its accuracy and completeness. All authors had final responsibility for the decision to submit for publication.

## Data Sharing Statement

The study protocol is provided as *Supplementary Appendix 2*. Individual participant (de-identified) data will be made available when the trial is complete upon direct request to the corresponding author with an appropriate research proposal. Once such a proposal is approved data will be shared through a secure online platform.

