## Supplementary for "Efficacy, safety, and lot to lot immunogenicity of an inactivated SARS-CoV-2 vaccine (BBV152): a, double-blind, randomised, controlled phase 3 trial"

### Supplementary Materials

**Table 1.** The COVAXIN Study Group **page 2**

Reverse transcription polymerase chain reaction (RT-PCR) **page 5**

Enzyme-linked immunosorbent assay (ELISA) at Screening **page 6**

Enzyme-linked immunosorbent assay (ELISA) for lot-to-lot comparisons **page 6**

Next-Generation Sequencing method **page 7**

Microneutralisation method **page 8**

ELISA Assays for S1 protein, RBD and N-antigen of SARS-COV-2 **page 9**

**Table 2.** Participants disposition status **page 10**

**Figure 1.** Geometric mean ratios of $MNT_{50}$ for three consecutive vaccine lots **page 11**

**Table 3.** Geometric mean $MNT_{50}$ by age, gender and baseline serostatus **page 12**

**Table 4.** Summary of adverse events **page 13**

**Table 5.** Incidences of solicited adverse events after each dose **page 14**

| **Supplementary table 1:** COVAXIN Study Group and Ethic Committees from all participating trial sites | | | | |
|---|---|---|---|---|
| **Hospital Name, City, State** | **Ethics Committee Number** | **Principal Investigator** | **Co-Principal Investigator** | **Study Coordinators/Field Worker/Nurse** |
| Lokmanya Tilak Municipal Medical College and General Hospital, Mumbai, Maharashtra | ECR/175/Inst/MH/2013/RR-19 | Nilkanth Awad, MD | 1) Siddharth Waghmare, MD<br>2) Vaibhav Aglawe, MD<br>3) Jairaj Nair, MD | 1) Bhavika Jain<br>2) Chaitali Babar<br>3) Poonam Kabale<br>4) Akash Dhoble<br>5) Ankur Humare |
| Grant Government Medical College and Sir J.J Group of Hospitals, Mumbai, Maharashtra | ECR/382/Inst/MH/2013/RR-19 | Priti Meshram, MD | 1) Dinesh Dhodi, MD<br>2) Manali Vable, MD<br>3) Archana Bandkar, MD | 1) Mehul Shah<br>2) Zainab Shaikh<br>3) Sabir Khan<br>4) Alex Pandit<br>5) Smruti Chalke |
| People's College of Medical Sciences and Research Centre And Associated People's Hospital, Bhopal, Madhya Pradesh | ECR/519/Inst/MP/2014/RR-17 | Raghvendra Gumashta, MD | 1) Sushil Jindal, MD<br>2) Chittaranjan Chaubal, MD<br>3) Sanjay Tandon, MD | 1) Saroj Mani<br>2) Anjlika Jhariya<br>3) Namrata Manjhi<br>4) Richa Jain |
| Nizam's Institute of Medical Sciences, Punjagutta Market, Hyderabad | ECR/303/INST/AP/2013/RR-19 | Prabhakar Reddy, MD | 1) Ruby Raphael, MD<br>2) V Bhavani, MD<br>3) Abid Ali, MD | 1) G. Devika<br>2) Lanka Tejaswi |
| Gujarat Medical Education and Research Society Medical College and Civil Hospital, Sola, Ahmedabad, Gujarat | ECR/404/Inst/GJ/2013/RR-20 | Parul Bhatt, MD | 1) Kiran Rami, MD<br>2) Rashmi Sharma, MD<br>3) Meera Shah, MD | 1) Smruti Parekh<br>2) Vaishali Chauhan<br>3) Viraj Salvi<br>4) Anuj Tarpara<br>5) Dhaval Kumpavat |
| Indian Council of Medical Research, National Institute of Cholera and Enteric Diseases, Kolkata, West Bengal | ECR/416/Inst/WB/2013/RR-20 | Suman Kanungo, MD | 1) Shanta Dutta, MD<br>2) Agniva Majumdar, MD<br>3) Jaayanta Saha, MBBS | 1) Snehasish Saha<br>2) Rupesh Mishra<br>3) Abhijit Guha<br>4) Dipak Das<br>5) Arpan Mitra |
| Institute of Medical Sciences and SUM Hospital, Bhubaneshwar, Odisha | ECR/627/Inst/OR/2014/RR-17 | Venkat E Rao, MD | 1) Jyotiranjan Sahoo, MD<br>2) Smaraki Mohanty, MD<br>3) Sandeep Kumar Panigrahi, MD | 1) Sahazad Ali<br>2) Banajini Pradhan<br>3) Manini Sahoo<br>4) Perween Sultana<br>5) Binata Samal |
| All India Institute of Medical Sciences, Patna, Bihar | ECR/1387/INST/BR/2020 | Chandramani Singh, MD | 1) Sanjay Pandey, MD<br>2) Pragya Kumar, MD<br>3) Yogesh Kumar, MD | 1) Sarvesh Singh<br>2) Nitin Kr Singh<br>3) Achal Singh<br>4) Shreekant Kumar |
| Jeevan Rekha Hospital, Belgaum, Karnataka | ECR/1242/INST/KA/2019 | Amit Bhate, MD | 1) Suresh Bhate, MD<br>2) Paritosh Desai, MD<br>3) Abhishek Chavan, MD | 1) Ajay Kunal<br>2) Imran Mulla<br>3) Sameer Nadaf<br>4) Laxman Arabhavi<br>5) Shoiab Shaikh |
| SRM Medical College and Research Centre, SRM | ECR/431/INST/TL/2013/RR-19 | Satyajit Mohapatra, MD | 1) Melvin George, MD<br>2) Balaji Ramraj, MD | 1) Tharunya Palanivel<br>2) Kalai Selvi<br>3) Indiraa Priyadarshini |

| | | | | |
|---|---|---|---|---|
| Nagar, Kattankulathur, Tamilnadu | | | 3) Gayathri Balasubramaniyam, MD | 4) Kamatchi<br>5) Anusuya |
| All India Institute of Medical Sciences, Patna, New Delhi | ECR/547/INST/DL/2014/RR-17 | Sanjay Rai, MD | 1) Randeep Guleria, MD<br>2) Shashi Kant, MD<br>3) Praveen Aggarwal, MD | 1) Shreya Jha,<br>2) Suprakash Mandal<br>3) Tripti Rai<br>4) Priyanka Bansal<br>5) Akshay Sharma<br>6) Kirti Diswar |
| Prakhar Hospital, Kanpur, Uttar Pradesh | ECR/1017/INST/UP/2017 | Jitendra Singh, MD | 1) V Tripathi, MD<br>2) Vikas Mishra, MD<br>3) Anit Singh, MD | 1) Nidhi Singh<br>2) Astha Singh<br>3) Saumya Singh<br>4) Sandeep Uniyal<br>5) Dev Chaudhary |
| Rahate Surgical Hospital and ICU, Nagpur, Maharashtra | ECR/601/Inst/MH/2014/RR-17 | Manish Multani, MD | 1) Prashant Rahate, MS | 1) Ashish Tajne<br>2) Vaishali Tajne<br>3) Vrushali Mohitkar<br>4) Pravina Lanjewar<br>5) Megha Balbudhe |
| Vydehi Institute of Medical Sciences and Research, Bengaluru, Karnataka | ECR/747/Inst/KA/2015/RR-18 | Akshatha Savith, MD | 1) Gummadi Reddy, MD<br>2) Chalapathy DV, MD<br>3) Natesh Rao, MD | 1) Nripesh Nepal<br>2) Khushboo Sharma<br>3) Hasina Taj<br>4) Arun Kumar<br>5) Payel Sarkar |
| Mahatma Gandhi Medical College and Research Institute, Pondicherry, Tamilnadu | ECR/451/Inst/PO/2013/RR-19 | Pajanivel Ranganadin, MD | 1) Lokesh Shanmugam, MD<br>2) Vimal Raj, MD | 1) Kowshik Reddy<br>2) Subashri R<br>3) Soundariya<br>4) Selva Pandian<br>5) Agnus Panikar |
| Redkar Hospital and Research Centre, Pernem, Goa | ECR/902/INST/GA/2018 | Sagar Redkar, MD | 1) Vivek Redkar, MD<br>2) Supriya Redkar, MD<br>3) Shraddha Rane, MD | 1) Tejaswini Patil<br>2) Mrunali Desai<br>3) Sarvesh Kerkar<br>4) Jyoti Patil<br>5) Dhananjay Lad |
| ESIC Medical College and Hospital, Haryana | ECR/167/Inst/HR/2013/RR-19 | Anil Pandey, MD | 1) Pooja Goyal, MD<br>2) Nidhi Anand, MBBS<br>3) Kranti Garg, MD | 1) Dr. Bharti Gaur<br>2) Neha Katiyar<br>3) Soniya Chahal<br>4) Ayona James<br>5) Mohit Prajapati |
| Directorate of Public Health and Preventive Medicine, Chennai, Tamilnadu | ECR/270/Inst/TN/2013/RR-16 | Selvavinayagam Sivaprakasam, MD | 1) Palani Sampath, MBBS<br>2) Sudharshini Subramaniam, MD | 1) Savitha Balaji<br>2) Roshini Azhaguvel<br>3) Parasuraman Palanivel<br>4) Sangliraj Muthukalai<br>5) Ayesha Bee |
| Sir Ganga Ram Hospital, New Delhi, Delhi | ECR/20/INST/DL/2013/RR-19 | Anupam Sachdeva, MD | 1) Manas Kalra, MD<br>2) Pooja Khosla, MD | 1) Ajeet Nanda<br>2) Vinay Sharma<br>3) Rajni Singh<br>4) Sakshi Pandey<br>5) Priyanka Singh |
| Maharaja Agrasen Hospital, Jaipur, Rajasthan | ECR/745/INST/DL/2015/RR-18 | Manish Jain, MD | 1) Prabhat Sharma, MD<br>2) Deepak Sharma, MBBS | 1) Kapil Soni<br>2) Khushwant Khatri |

| | | | | |
|---|---|---|---|---|
| | | | 3) Madhvender Jain, MD | 3) Sanjeev Vimal<br>4) Gaurav Dalvi<br>5) Preshita Vanjare |
| Pandit Bhagwat Dayal Sharma Post Graduate Institute of Medical Sciences, Rohtak, Haryana | ECR/293/Inst/HR/2013/RR-19 | Savita Verma, MD | 1) Dhruva Chaudhary, MD<br>2) Ramesh Verma, MD<br>3) Pawan Singh, MD | 1) Deepak Gill<br>2) Anjali Ahlawat<br>3) Kavita<br>4) Bijender Singh<br>5) Rosy |
| Government Fever Hospital, Gorantla, Guntur | ECR/467/Inst/AP/2013/RR-19 | S Laxmikumari, MD | 1) D Sudheer, MD<br>2) P Basha, MD | 1) Krishna Suri<br>2) Durga Anjali<br>3) G Divya<br>4) K Sowmya<br>5) K Jyothi |
| Aligarh Muslim University, Aligarh, Uttar Pradesh | ECR/1418/ Inst/UP/2020 | Mohammad Shameem, MD | 1) Mansoor Tariq, MS | 1) Mohammad Shiraz<br>2) Azimuddin Malik<br>3) Shafeequr Rahman<br>4) Nafees Khan<br>5) Samia Kirmani |
| Prakash lnstitute of Medical Science and Research, Islampur, Sangli, Maharashtra | ECR/1052/Inst/ MH/2018 | Vijaykumar Patil, MD | 1) Pradeep Kulkarni, MBBS<br>4) B Patil, MD | 1) Vishwajit Khade<br>2) Heyeshi Singh<br>3) Suhash Thorat |
| Rajashree Chhatrapati Shahu Maharaj Government medical college and Chhatrapati Pramila Raje Hospital, Kolhapur, Maharashtra | ECR/703/ Inst/MH/2015/RR-20 | Sunita Ramanand, MD | 1) Vijay Barge, MD<br>2) Varun Bafna, MD<br>3) Rama Bhosale, MD | 1) Ratnadeep Patil<br>2) Shivani Patil<br>3) Suraj Shewale<br>4) Sadhana Jadhav<br>5) Nupur Shevale |
| **Bharat Biotech Clinical Team:**<br>Shashi Kanth Muni, BDS<br>Sapan Kumar Behera, MD<br>Yuvraj Jogdand, MD<br>Bhargav Reddy Dalta, PharmD<br>Mr. Sunil Kumar Kantheti<br>Ms. Sandhya Rani Nandala<br>Ms. Aparna Bathula<br>Ms. Amaravani Pittala<br>Mr. Little Master Reddy<br>Mr. Ashok Sudamala<br>Mr. Nagaraju Pillutla<br>Mr. Hrishikesh Reddy<br>Ms. Akhila Naidu | | | **ICMR-National Institute of Virology:**<br>Anita Shete-Aich, PhD<br>Gururaj Deshpande, PhD<br>Dr. Vinita Malik, PhD<br>Mrs. Sheetal Kadam, MSc<br>Prof. (Dr.) Priya Abraham, MD, PhD<br><br>**Indian Council of Medical Research:**<br>Samiran Panda, PhD<br>Swati Gupta, PhD | |

**Reverse transcription polymerase chain reaction (RT-PCR)**

The ICMR-NIV 2019-nCoV Assay Kit 3.1 contains a set of TaqMan RT-PCR assay for the qualitative detection and characterisation of SARS-CoV-2 RNA. The assay kit uses TaqMan Fluorogenic probe-based chemistry that uses the 5' nuclease activity of Taq DNA polymerase and enables the detection of a specific PCR product as it accumulates during PCR cycles.

The assay includes three targets E gene, ORF 1ab, RdRp of SARS-CoV-2 genes, and one house keeping gene, the β-Actin gene. The assay has one screening and two confirmatory viral genomic region targets, reducing the risk of false negatives.

The assay runs for 40 cycles; however, for any interpretation, the threshold cut off cycle Ct is 35.

**Results Interpretation is as follows:**

| Target | E | ORF | RdRp | β-Actin |
|---|---|---|---|---|
| Positive | + | + | +/- | + |
| Negative | - | - | - | + |
| Strong* Positive (Very low Ct) | + | + | + | - |
| Sample quality poor | - | - | - | - |
| Inconclusive | + | - | - | + |

**Enzyme-linked immunosorbent assay (ELISA) at Screening**

The National Institute of Virology (NIV) SARS-CoV-2 Human IgG ELISA kit is intended for qualitative detection of IgG antibodies in serum/plasma of patients presenting clinical signs and symptoms consistent with SARS CoV-2 infection or recovered patients. The sensitivity of the assay is 97·9% percent and specificity 92·3% percent for Laboratory routine testing.

Each kit contains one vial of "Positive control" and one vial of "Negative control". These work as markers of kit performance. P/N ratio of Positive control is defined as ratio of OD value of Positive control divided by OD of average OD of Negative control. The test is considered to be valid if P/N ratio is greater than 1.5.

Results are interpreted as follows:

- For an unknown sample (test sample) if O.D. value > Cutoff value and P/N ratio more than 1.5, sample should be considered as "Positive".
- For an unknown sample (test sample) if O.D. value < Cutoff value and P/N ratio less than 1.5, sample should be considered as "Negative".

**Enzyme-linked immunosorbent assays (ELISA) done at Bharat Biotech for lot-to-lot comparisons**

ELISA tests were performed as per standard protocols. Briefly, microtiter plates were coated with SARS-CoV-2 specific antigens: Whole inactivated SARS CoV-2 antigen; spike (S1) (Syngene, Bangalore, India, Batch No# PRB026913); Receptor Binding Domain (RBD) (Syngene, Bangalore, India, Batch No#PRB025485); nucleocapsid (N) (Syngene, Bangalore, India, Batch No# PRB025627) at a concentration of 1µg/ml, 100µl/well in PBS pH 7·4). After overnight incubation, wells were blocked and serially diluted sera added. After incubation, goat anti-Human IgG HRP conjugate (Sigma-Aldrich, Cat# A8667, dilution 1:5000) was added and incubated for 1 hr at RT. Tetramethyl benzidine was used as a substrate and absorbance measured at 450/630nm. Threshold value (Mean + 3 SD) was established by taking the absorbance of Day 0 sera and antigen-specific endpoint titers were determined for Day 56 sera samples. The reciprocal antibody dilution, at which absorbance is above the threshold, was taken as antigen-specific antibody endpoint titers. All methods were validated with respect to sensitivity and specificity.

Known unvaccinated and uninfected individual sera were used as a negative controls. Simultaneously, ELISA blank (without coating antigen) was also maintained as a negative control. Apart from this, cut off (Mean+3 SD) was drawn from the absorbance obtained at various dilutions (1:1000 to 1:32000) of sera collected on day 0 (before vaccination) which had been found negative in the RT-PCR and serology tests.

**Next-Generation Sequencing**

We were able to collect additional Nasopharyngeal swabs (NP) swabs from symptomatic Covid-19 (diagnosed by RT-PCR) participants who provided additional consent. All of the sequences were generated by the ICMR-National Institute of Virology (NIV), Pune, India using a next-generation sequencing approach. Controls were checked to ensure no evidence of amplification in the negative tests and that expected RNA quantification was consistent with cycle threshold (Ct) values provided by the testing laboratories.

All samples were processed by NIV laboratory staffs who were masked to vaccine allocation. In brief, the total RNA was extracted from 200-400 µl of the SARS-CoV-2 real-time RT-PCR positive samples. Extracted RNA was quantified using a Qubit RNA High Sensitivity (HS) kit by Qubit® 2.0 Fluorometer (Invitrogen, Life Technologies). Samples having CT values below 25 were selected for further processing with NGS. A total of 208 primers (104 for Pool#1 and 104 for Pool #2) were designed using the Primal Scheme online tool covering the entire genome of the SARS-CoV-2. A multiplex RT-PCR using the Superscript IV PCR kit (Invitrogen) was performed under the following conditions: 50°C for 30 min; 98°C: 2 min; 35 cycles of 98°C for 10 sec, 56°C for 100 sec, and 72°C for 30 sec and final extension at 72°C for 5 min. The PCR products obtained were loaded (25 µl) onto a 2% Agarose gel and checked for the presence of a PCR product of the desired size.

Bands of around 400 bp were observed in all the Pool 1 and Pool 2 PCR products of samples. Gel extraction of DNA bands was performed using the QIA quick DNA extraction kit protocol. This gel-purified DNA product was used for the library preparation from the A-tailing step by Illumina Truseq LT kit as described earlier (Nyayanit et al, 2020; Yadav et al, 2021). Libraries were quantified using KAPA Library Quantification Kit (Kapa Biosystems, Roche Diagnostics Corporation, USA). Equimolar ratios of libraries were pooled and denatured with 0·1 N NaOH and were neutralised using 0·1 M Tris (pH7·0). The denatured libraries were diluted to 1·3 picomole using hybridization buffers before loading onto Illumina Next seq 550 mid-outputs 300 cycles' reagent cartridges. CLC genomics workbench version11.0 (CLC, QIAGEN, and Germany) was used for the analysis of the data generated from the machine. Reference-based mapping was performed to retrieve the sequence of the SARS-CoV-2.

*Nyayanit DA, Sarkale P, Baradkar S, et al. Transcriptome & viral growth analysis of SARS-CoV-2-infected Vero CCL-81 cells. Indian J Med Res 2020; 152:70–6.*

*Yadav PD, Potdar VA, Choudhary ML, et al. Full-genome sequences of the first two SARS-CoV-2 viruses from India. Indian J Med Res 2020; 151: 200–9.*

**Microneutralisation immunogenicity assay method ($MNT_{50}$)**

A microneutralisation assay ($MNT_{50}$) was done at Bharat Biotech for the immunological lot to lot comparisons.

The sera collected from all enrolled participants were inactivated at 56°C in a water bath for 30 min. Sera were successively diluted in a two-fold series from a starting dilution of 1:8 to the required concentration, and an equal volume of challenge virus solution containing 100 $CCID_{50}$ viruses was added. After neutralisation in a 37°C incubator for two hours, a 1.0 x $10^5$ /mL cell suspension was added to the wells (0.1 mL/well) and cultured in a $CO_2$ incubator at 37°C for 3–5 days. The method of Ramakrishnan (2016) was used applied to observations of the cytopathic effect (CPE) to calculate the neutralisation endpoint ($MNT_{50}$) converting to logarithm the serum dilution that protects 50% of cells from infection by challenge with 100 $CCID_{50}$ virus.

During each assay, a known antibody titre is used as a positive control, and pre-immune sera are used as a negative control.

*Ramakrishnan MA. Determination of 50% endpoint titer using a simple formula. World J Virol 2016; 5:85–6.*

**ELISA assays for IgG against SARS-Cov-2 epitopes**

The National Institute of Virology (NIV) SARS-CoV-2 Human IgG ELISA kit is intended for qualitative detection of IgG antibodies in serum/plasma of patients presenting clinical signs and symptoms consistent with SARS CoV-2 infection or recovered patients. The sensitivity of the assay is 97·9% percent and specificity 92·3% percent for Laboratory routine testing.

Each kit contains one vial of "Positive control" and one vial of "Negative control". These work as markers of kit performance. P/N ratio of Positive control is defined as ratio of OD value of Positive control divided by OD of average OD of Negative control. The test is considered to be valid if P/N ratio is greater than 1.5.

Results are interpreted as follows:

- For an unknown sample (test sample) if O.D. value > Cutoff value and P/N ration more than 1.5, sample should be considered as "Positive".
- For an unknown sample (test sample) if O.D. value < Cutoff value and P/N ration less than 1.5, sample should be considered as "Negative".

**Enzyme-linked immunosorbent assays (ELISA) done at Bharat Biotech for lot-to-lot comparisons**

ELISA tests were performed as per standard protocols. Briefly, microtiter plates were coated with SARS-CoV-2 specific antigens: Whole inactivated SARS CoV-2 antigen; spike (S1) (Syngene, Bangalore, India, Batch No# PRB026913); Receptor Binding Domain (RBD) (Syngene, Bangalore, India, Batch No#PRB025485); nucleocapsid (N) (Syngene, Bangalore, India, Batch No# PRB025627) at a concentration of 1µg/ml, 100µl/well in PBS pH 7·4). After overnight incubation, wells were blocked and serially diluted sera added. After incubation, goat anti-Human IgG HRP conjugate (Sigma-Aldrich, Cat# A8667, dilution 1:5000) was added and incubated for 1 hr at RT. Tetramethyl benzidine was used as a substrate and absorbance measured at 450/630nm. Threshold value (Mean + 3 SD) was established by taking the absorbance of Day 0 sera and antigen-specific endpoint titers were determined for Day 56 sera samples. The reciprocal antibody dilution, at which absorbance is above the threshold, was taken as antigen-specific antibody endpoint titers. All methods were validated with respect to sensitivity and specificity.

Known unvaccinated and uninfected individual sera were used as a negative controls. Simultaneously, ELISA blank (without coating antigen) was also maintained as a negative control. Apart from this, cut off (Mean+3 SD) was drawn from the absorbance obtained at various dilutions (1:1000 to 1:32000) of sera collected on day 0 (before vaccination) which had been found negative in the RT-PCR and serology tests.

| **Supplementary table 2:** Participants disposition status | | | |
|---|---|---|---|
| **Description** | **BBV152 (N=12,899) n (%)** | **Placebo (N=12,899) n (%)** | **Total (N=25,798) n (%)** |
| Screened Participants | | | 26028 |
| Screen Failure | | | 217 |
| Screened but not enrolled | | | 13 |
| Enrolled Participants | 12899 (100) | 12899 (100) | 25798 (100) |
| Ongoing Participants | 12194 (94·5) | 12063 (93·5) | 24257 (94·.0) |
| Randomisation Analysis Set [a] | 12899 (100) | 12899 (100) | 25798 (100) |
| Full Analysis Set (FAS)[b] | 8946 (69·4) | 8988 (69·7) | 17934 (69·5) |
| Per-protocol Analysis Set (PP)[c] | 8471 (65·7) | 8502 (65·9) | 16973 (65·8) |
| Immunogenicity Analysis Subset [d] | 428 (3.3) | 141 (1.1) | 569 (2·2) |
| Safety Analysis Set [e] | 12879 (99·8) | 12874 (99·8) | 25753 (99·8) |
| Category of Site | | | |
| Category 1 (Symptomatic) | 8102 (62·8) | 8375 (64·9) | 16477 (63·9) |
| Category (Symptomatic/Asymptomatic) | 4348 (33·7) | 4373 (33·9) | 8721 (33·8) |
| Category 3 (Symptomatic/Asymptomatic + Immunogenicity) | 449 (3·5) | 151 (1·2) | 600 (2·3) |
| Visit 1 Completed | 12,899 (100) | 12,899 (100) | 25,798 (100) |
| Visit 2 Completed | 12,310 (95·4) | 12,310 (95·4) | 24,620 (95·4) |
| Visit 3 Completed | 12,054 (93·5) | 12,086 (93·7) | 24,140 (93·6) |
| Visit 4 Completed | 11,909 (92·3) | 11,893 (92·2) | 23,803 (92·3) |

(a) All randomised participants classified according to the study product (vaccine or placebo) to which they were randomised.

(b) All randomised participants who received at least one dose of IP and had no immunologic evidence of prior COVID-19 (i.e, negative against SARS-CoV-2 antibodies) at Visit 1 before the first dose of IP. Participants will be analysed according to the study product (vaccine or placebo) received.

(c) All participants in the FAS who received planned doses of IP per schedule, seronegative for SARS-CoV-2 antibody by ELISA at baseline, and had no major protocol deviations, as determined, and documented by the Sponsor.

(d) Designated participants at study sites included in the immunogenicity study (category 3) who had received both doses of IP and had no major protocol deviations.

(e) The Safety Set consists of all randomised participants who received at least one dose of IP, classified according to the study product received.

% = n/N *100 where N = number of enrolled participants and n = number of participants.

Database cut-off date: 17 May 2021

**Figure 1.** Geometric mean ratios of $MNT_{50}$ for three consecutive manufacturing lots of BBV152

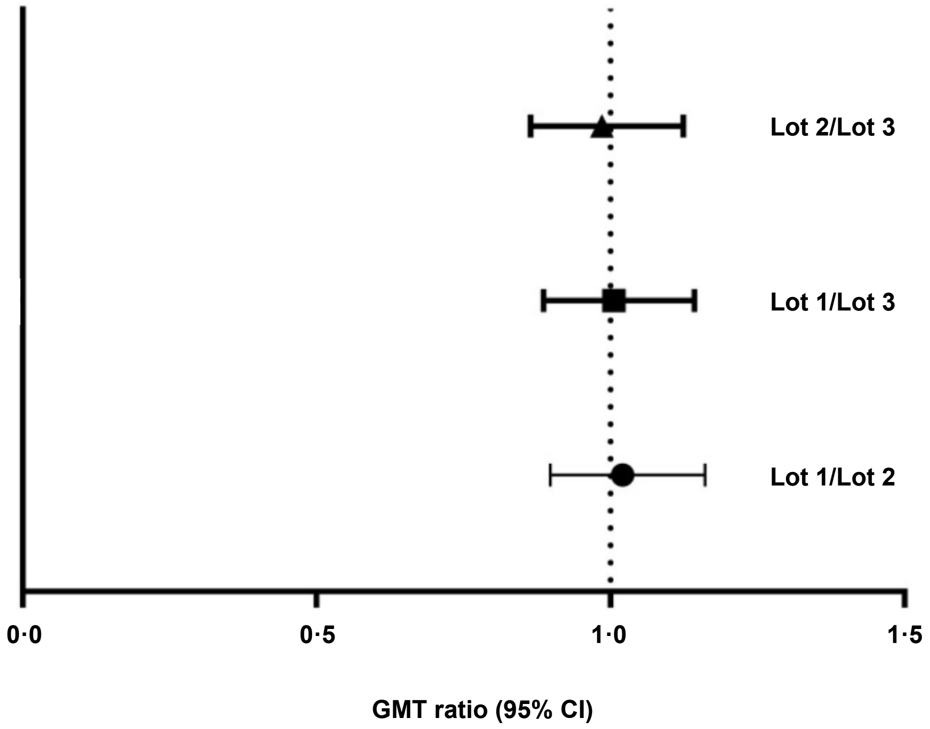


Lot-to-lot consistency between treatments was declared if for all lot-to-lot comparisons, the two-sided 95% CI for the GMC ratio was completely contained within 0·5 and 2·0.

**Supplementary table 3:** $MNT_{50}$ neutralising antibody titres (95% CI) at Day 56 by age and gender

| **$MNT_{50}$** | | **BBV152 (All lots) (N = 386)** | | **Placebo (N = 119)** | |
|---|---|---|---|---|---|
| | | n | GMT (95% CI) | n | GMT (95% CI) |
| **Age Group (years)** | **≥18–<60** | 334 | 129.9 (114.3, 147.6) | 101 | 12.9 (10.1, 16.5) |
| | **≤60** | 52 | 101.2 (70.0, 146.3) | 18 | 19.1 (9.0, 40.5) |
| **Gender** | **Male** | 238 | 118.2 (101.0, 138.3) | 77 | 14.1 (10.4, 19.2) |
| | **Female** | 148 | 138.4 (114.4, 167.3) | 42 | 12.9 (8.8,19.0) |
| **SARS-CoV-2 IgG status at baseline** | **Negative** | 338 | 118.03 (104.0, 134.0) | 99 | 11.9 (9.3,15.2) |
| | **Positive** | 48 | 194.3 (134.4, 280.9) | 20 | 27.4 (14.0, 53.5) |

Shown are geometric mean titres measured using the wild-type SARS-CoV-2 microneutralisation assay ($MNT_{50}$) in sera obtained at Day 56, 4 weeks after the second vaccination.

**Supplementary table 4.** Summary of adverse events

| | BBV152 (N = 12,879) | | Placebo (N = 12,874) | | Total (N = 25,798) | |
|---|---|---|---|---|---|---|
| | Events n | Participants n (%) | Events n | Participants n (%) | Events n | Participants n (%) |
| **All Adverse Events** | 2930 | 1597 (12·4) | 3029 | 1597 (12·4) | 5959 | 3194 (12·4) |
| **Solicited AEs** | | | | | | |
| Any solicited AE | 1949 | 1223 (9·5) | 1720 | 1136 (8·8) | 3669 | 2359 (9·2) |
| Solicited AE within 7 days post dose 1 | 1151 | 809 (6·3) | 994 | 702 (5·5) | 2145 | 151 (5·9) |
| Solicited AE within 7 days post dose 2 | 798 | 568 (4·4) | 726 | 548 (4·3) | 1524 | 1116 (4·3) |
| **Unsolicited AEs** | 981 | 489 (3·8) | 1309 | 609 (4·7) | 2290 | 1098 (4·3) |
| **Serious Adverse Events** | 40 | 39 (0·30) | 66 | 60 (0·47) | 106 | 99 (0·38) |
| **All Medically Attended Adverse Events (MAAEs)** | 475 | 301 (2·3) | 548 | 319 (2·5) | 1023 | 620 (2·4) |
| **Immediate AEs** (within 30 min post vaccination) | | | | | | |
| Any immediate AE | 14 | 12 (0·10) | 29 | 23 (0·18) | 43 | 35 (0·14) |
| Immediate AEs post dose 1 | 11 | 10 (0·08) | 19 | 17 (0·13) | 30 | 27 (0·10) |
| Immediate AEs post dose 2 | 3 | 3 (0·02) | 10 | 8 (0·06) | 13 | 11 (0·04) |
| **All Adverse Events of Special Interest (AESI)** | 23 | 23 (0·18) | 23 | 23 (0·18) | 46 | 46 (0·18) |

N = number of participants in the relevant population,
Events, n = number of individual events reported (one participant may have reported several AEs),
Participants, n = Number of participants reporting at least one event,
% = n participants with an event/N *100

**Supplementary table 5.** Incidences of solicited adverse events after each dose.

| **Participants reporting solicited AEs within 7 days of vaccination**, n(%) | **BBV152** (N = 12,879) | | **Placebo** (N = 12,874) | | **Total** (N = 25,798) | |
|---|---|---|---|---|---|---|
| | Dose 1 n (%) | Dose 2 n (%) | Dose 1 n (%) | Dose 2 n (%) | Dose 1 n (%) | Dose 2 n (%) |
| **Any Local AE** | 431 (3·35) | 278 (2·16) | 399 (3·10) | 260 (2·02) | 830 (3·22) | 538 (2·09 |
| Pain | 392 (3·04) | 233 (1·81) | 358 (2·78) | 208 (1·62) | 750 (2·91) | 441 (1·71) |
| Erythema | 33 (0·26) | 21 (0·16) | 26 (0·20) | 25 (0·19) | 59 (0·23) | 46 (0·18) |
| Induration | 32 (0·25) | 18 (0·14) | 26 (0·20) | 18 (0·14) | 58 (0·23) | 36 (0·14) |
| Swelling | 21 (0·16) | 14 (0·11) | 32 (0·25) | 16 (0·12) | 53 (0·21) | 30 (0·12) |
| **Any Systemic AE** | 331 (2·57) | 231 (1·79) | 247 (1·92) | 205 (1·59) | 578 (2·24) | 436 (1·69) |
| Pyrexia | 108 (0·84) | 86 (0·67) | 81 (0·63) | 79 (0·61) | 189 (0·73) | 165 (0·64) |
| Fatigue | 52 (0·40) | 41 (0·32) | 41 (0·32) | 20 (0·16) | 93 (0·36) | 61 (0·24) |
| Chills | 28 (0·22) | 9 (0·07) | 22 (0·17) | 16 (0·12) | 50 (0·19) | 25 (0·10) |
| Headache | 128 (0·99) | 86 (0·67) | 111 (0·86) | 70 (0·54) | 239 (0·93) | 156 (0·61) |
| Myalgia | 49 (0·38) | 37 (0·29) | 28 (0·22) | 28 (0·22) | 77 (0·30) | 65 (0·25) |
| Arthralgia | 17 (0·13) | 12 (0·09) | 17 (0·13) | 17 (0·13) | 34 (0·13) | 29 (0·11) |
| Nausea | 17 (0·13) | 14 (0·11) | 12 (0·09) | 10 (0·08) | 29 (0·11) | 24 (0·09) |
| Vomiting | 12 (0·09) | 6 (0·05) | 8 (0·06) | 8 (0·06) | 20 (0·08) | 14 (0·05) |

N = number of participants in the relevant population,
Events, n = number of individual events reported (one participant may have reported several AEs),
Participants, n = Number of participants reporting at least one event,
% = n participants with an event/N*100

Med
